# “An explosion of alternative medicines in France!”: media-biased polyphonic epidemiology vs. evidence-based data

**DOI:** 10.1101/2023.09.03.23294995

**Authors:** Fabrice Berna, Nans Florens, Laurence Verneuil, François Paille, Chantal Berna, Julien Nizard

## Abstract

**Background:** The media and several French official organizations report an “explosion” in the use of complementary and alternative medicines (CAM) in France, possibly “boosted” by the Covid-19 pandemic. In addition, the French Miviludes (Mission of Vigilance and Fight Against Sectarian Aberrations) routinely asserts in its yearly reports “a dramatic uptick” in sectarian aberrations in the health sector. However, data supporting those assertions are either scant or rarely compared with data from previous years to properly apprehend an evolution.

This paper aims to analyze existing data in this field and to examine in which domain (population-based survey, official reports, media) “increases” or “explosion” might be observed. We compared the data from France and Switzerland. In fact, no clear regulatory policy on CAM exists in France, whereas the Swiss population voted to include CAM into the Swiss Constitution in 2009.

**Method:** Surveys documenting the reported usage of CAM in both France and Switzerland were analyzed. Health-related sectarian aberrations were analyzed based on yearly reports of the French Miviludes and its Swiss counterpart (Inter-Cantonal Information Center on Beliefs). Then, the communication in the media on CAM was analyzed in the French media and in the scientific literature published in both countries. Three time periods were considered to apprehend the claimed boost of the Covid-19 pandemic.

**Results:** Our comprehensive analysis of available information sources does not suggest an “explosion” in CAM use or related sectarian misconduct. Reported CAM use in France was 39% in 2007 and later surveys did not find higher percentage. Reported CAM use increased from 24% to 28.9% between 2007 and 2017 in Switzerland. Referrals about health-related sectarian aberrations regularly increased until 2017 and then remained stable in France; they regularly decreased in Switzerland. Evidence for a pandemic boost was weak. In contrast, a steady increase was observed in the number of mentions in media of CAM.

**Conclusion:** Our analyses indicate a media-skewed, polyphonic epidemiology, which is not supported by available data. Health care specialists, Official organizations, journalists and politicians should become aware of biases concerning CAM and foster a more informed and balanced discourse regarding these practices.

## 1. Background

Media affect the way we perceive the world around us, with some interesting contradictions. For example, a worrying increase of “irrational beliefs about pseudo-sciences” was recently reported in a survey conducted on French youths [1]. It claims that “scientific misinformation” has dramatically increased following the Covid-19 pandemic, pointing the responsibility to social networks. However, a careful analysis of methodological caveats rather discarded both this claimed “increase of irrational beliefs” and the causal influence of “scientific misinformation” relayed by social networks (Evrard R., manuscript in review).

In the field of “complementary and alternative medicines”, a similar phenomenon is observed in France where several surveys, but also official institutions, have reported a significant increase in the use of UnConventional Healthcare Practices (UCHP, the term for “complementary and alternative medicines” chosen by the French Ministry of Health and Prevention). Terms like “essor,” “exponential increase,” and “explosion” are thus used to describe this phenomenon [2]. The French Ministry of Health and Prevention has even established a technical support committee to oversee UCHP, which was launched in June 2023 under the authority of the Minister Delegate for Territorial Organization and Health Professionals[3]. The use of terms like “exponential” and “explosion” suggests that the phenomenon is perceived as being “out of control”, particularly since the Covid-19 pandemic, which has further fueled mistrust of conventional medicine (see [4], [5] p.187).

An increased use of UCHP has been broadly reported in French-language media. However, the portrayal of UCHP in the media rarely remains neutral. Sectarian deviations are often highlighted. A partisan perspective leads to either extolling certain practices for their alleged virtues and promises, which are seldom backed by scientific proof, or emphasizing the potential risks tied to these practices, like loss of opportunity or harmful consequences, without recognizing any potential benefits. The term “alternative medicine”, frequently used in the media, perpetuates this confusion. It pertains to the application of UCHP rather than the practices themselves [6, 7]. For instance, the “alternative use” of UCHP as a replacement for effective conventional treatment in cancer cases inevitably leads to substantial risk of loss for patients [8]. However, this “alternative use” should not be mistaken for the use of UCHP in combination with conventional treatments, i.e., complementary or integrative medicine, as endorsed by the American Society for Clinical Oncology [9].

Likewise, the threat of sectarian deviations, often simplistically linked with certain UCHP [5], needs a more nuanced understanding, considering factors like the method utilized, the therapist’s training and therapeutic stance, the medical condition it’s intended for, and the context in which the UCHP is delivered (e.g., hospital, private practice, etc.). For example, suggesting mindfulness-based methods in a health-care setting should not be confused with Buddhistic meditation in a religious or spiritual context. It’s important to remember that no practice, unconventional or conventional medicine alike, is completely devoid of potential risks of therapeutic deviations.

Consequently, the Strengths, Weaknesses, Opportunities, and Threats (SWOT) analysis presented to the public appears incomplete or distorted, failing to fully encapsulate the complexity of UCHP [10].

Beyond the media, recent publications by French institutions also adhere to message about rapidly rising numbers [5, 11]. The latest report from the French National Medical Council [11] speaks of an “exponential increase” in the use of UCHP, though it fails to provide numeric data, whether first hand or as a citation. Instead, the report bolsters its claim by citing growing reports of deviations that are becoming “increasingly significant, and even occurring daily”. The media covering this publication [12], reported that about 1700 reports may have been received by the French National Medical Council in 2022. However, no comparative data from previous years was provided. Even by the section of the French Press Agency in charge of fact-checking reported an “explosion of UCHP (..) boosted by the unprecedented health system crisis” without providing any factual data [4]. The same phenomenon was observed in a previous official report sent to the French Prime Minister in 2012 on UCHP: a “growing interest” was already mentioned, with a single-time point prevalence of use in France [13]. This report also stated there was “an increase of UCHP around the world”, although available data at that time provides no support for it [14].

Another entity, the “Mission of Vigilance and Fight Against Sectarian Aberrations” (ie “Miviludes” standing for “Mission Interministérielle de Vigilance et de Lutte contre les Dérives Sectaires” in French), also routinely asserts in its reports “a dramatic uptick” in sectarian aberrations in the health sector. Miviludes, is a national agency established in 2003 and under the purview of the French Interior Ministry since 2020. It maintains a somewhat distinctive policy regarding cults and sectarian deviations in Europe [15]. Upon Miviludes’ inception, France took a leading position in Europe regarding its fight and pro-active policy against cults [16]. Nations such as Belgium, Poland and Russia have adopted similar policies [16], whereas others like Switzerland [17], Germany [18] and other West European nations opted for a weakly interventionist policy.

In this paper, our primary analysis will focus on France, given the recent establishment of the technical support committee for supervising UCHP [3]. Nevertheless, we will incorporate data from Switzerland, a neighboring country where the media also claim a similar “surge” in the use of UCHP [19]. Comparing the situation with Switzerland is especially pertinent as, in 2009, the population voted to include du UCHP into the Swiss Constitution. This led to the formal inclusion of “complementary and integrative medicines” into the medical faculty curriculum, as well as the appointment of chairs for Complementary and Integrative Medicine) at 3 of the 5 main medical schools [20]. Furthermore, Federal diploma for different UCHP have been established, including naturopathy, art therapy and massage therapy.

Interestingly, the “Inter-Cantonal Information Center on Beliefs” (CIC) was established in Switzerland in 2002, slightly before France’s similar initiative. The CIC is responsible for handling both health-related and religious aberrations. Unlike France’s Miviludes, the CIC is a private foundation of public utility, essentially a non-political organization composed of several academics and researchers. Its role is to offer neutral information and training courses to the public, functioning in a less proactive manner compared to its French counterpart.

This study aims to confront the data at hand regarding the evolution over the past decade of the use of UCHP in France and Switzerland by the population, with the UCHP-related misconduct cases as well as with mentions in the media of UCHP.

### 2. Method

#### 2.1. Sources

Three distinct topics were considered: one documenting the use of UCHP in the population, a second the UCHP-related aberrations and a third the communication in the media on UCHP.

The use of UCHP in the population was based on reviews of recent surveys conducted in France and Switzerland [14, 21–24].

UCHP-related misconducts were extracted from the yearly reports of the Miviludes and the CIC. Numerical data were available from 2010 to 2021 for Miviludes [25] and from 2003 to 2022 for CIC [26].

Regarding media communication on UCHP, we conducted two complementary analyses. The first analysis (Analysis 1, see details below), was centered on major French newspapers. This analysis documented the number of articles with a UCHP-related topics. A second bibliographical analysis (Analysis 2, see details below), examined the scientific literature published on this topic.
Analysis 1: we counted separately the number of articles published each year that included the following keywords: “acupuncture”, “homeopathy”, “osteopathy”, “naturopathy”, and “miviludes”. The web-thesaurus of each newspaper was used. Authors agreed in selecting 6 French newspapers among those most read and with a large scope of news in France (3 daily newspapers: Le Monde, Le Figaro, Libération, and 3 weekly newspapers: Le Point, L’Express, L’Obs). Unfortunately, we were not able to conduct the same analysis in Swiss journals, due to a limited access to the newspapers’ web-thesaurus.
Analysis 2. We counted the number of articles published each year on the PubMed plateform. We included those with the following keywords in their title or abstract: “complementary alternative medicine” OR “alternative medicine” OR “integrative medicine” OR “acupuncture” OR “homeopathy” OR “naturopathy” OR “osteopathy” OR “hypnosis”. A higher number of keywords was used as they reflect more how the scientific community labels UCHPs. The keyword “Miviludes” or “CIC” were not included as this committee is not known from the scientific field. As our analysis aimed to focus on France and Switzerland, articles published in English, French (for France), and in German or Italian (for Switzerland) were considered.

#### 2.2. Time periods

In order to gauge any “recent” and “exponential increase”, we evaluated changes in UCHP-related information across three distinct periods:

Period 1: From 2010 to the latest available data, with 2010 marking the initiation of the new UCHP regulatory policy in Switzerland and the availability of the first numerical data from Miviludes. It’s also a decade prior to the pandemic.
Period 2: The five years preceding the pandemic (2015-2016) up to the latest available data.
Period 3: The post-pandemic period (from 2020 up to the latest available data).
The latest data were from 2021 for Miviludes, 2022 for CIC, and June 30th, 2023 for newspapers and PubMed analyses. As a result, we projected by doubling the number of articles for 2023 to allow for a more effective comparison with prior years. We calculated the percentage increase or decrease of referrals (Miviludes, CIC) or articles (newspapers, PubMed) from the latest available data relative to the data at the onset of each time period. For each period, the value of onset was obtained by averaging the values of 2010-2011, 2015-2016, and 2018-2019. The value of the end of period was that of the last available data.

As data from surveys were not available for every year, their results are presented independently of those time periods.

### 3. Results

#### 3.1. A Surge in the Use of UCHP? What Do the Surveys Reveal?

In France, four surveys were available between 2007 and 2023 [21–24]. They were conducted on representative samples of 1,000 to 1,500 individuals. In Switzerland, three major surveys have been conducted on larger representative sample of about 15,000 individuals in 2007 and 2017. The summary results of these surveys are presented in Table S1 in supplementary material (SM).

In France, the 2007 survey revealed that 39% of respondents reported having used at least one type of “natural medicine” in the past year [21]. In 2019, 11 UCHPs were identified with usage in the past year ranging from 5.0% (CI95%:3.9-6.1) (for hypnosis) to 32.0% (CI95%:29.7-34.3) (for osteopathy) among those questioned [22]. In 2023 [24], 14 UCHPs were identified and the figure ranged from 7.5% (CI95%:6.1-8.9) (for kinesiology) to 35.9% (CI95%:33.1-38.7) (for osteopathy). Bearing in mind that comparing surveys should remain cautious, an increase in the reported use of homeopathy (from 17.8 (CI95%:18.0-19.6) to 30.7% (CI95%:27.9-33.5)), and hypnosis (4.5% (CI95%:3.4-6.6) vs. 10.2% (CI95%:8.7-11.7)) was observed between 2019 and 2023. In contrast, the reported use of osteopathy (31.9% (CI95%:29.6-34.3) vs. 35.9% (CI95%:33.1-38.7)) and acupuncture (12.0% (CI95%:10.5-13.5) vs. 11.2% (CI95%:9.7-12.7)) was stable.

Another 2021 survey demonstrated that the portion of respondents with a positive perception of UCHPs had risen from 80% (CI95%:78.0-82.0) in 2019 to 85% (CI95%:83.0-87.0) in 2021[23], and then dropped to 70% (CI95%:68.2-72.8) in 2023[24].

In Switzerland, reported usage of UCHPs at least once in the preceding year was stable between 2007 (24.0%, CI95%:23.1-25.0) and 2012 (24.7% CI95%:23.9-25.4), and then significantly increased in 2017 (28.9.0%, CI95%:28.1-29.7) [14, 27]. A significant increase was observed for homeopathy between 2007 (6.4%, CI95%:5.8-6.9) and 2012 (8.2%, CI95%:7.7-8.7), but not between 2012 and 2017 (8.4%, CI95%:8.0-8.9). Reported use of naturopathy was stable between 2007 (7.7%, CI95%:7.2-8.3) and 2012 (7.7%, CI95%:7.2-8.2) and then significantly increased in 2017 (8.8%, CI95%:8.3-9.0). Significant and constant increases were observed for osteopathy, and herbal medicine between 2007, 2012, and 2017 (osteopathy: 5.4%, CI95%:5.0-5.9 in 2007, 6.8%, CI95%:6.4-7.2 in 2012, 9.5%, CI95%:9.0-10.0 in 2017; herbal medicine: 2.7%, CI95%:6.4-7.2 in 2007, 5.0%, CI95%:6.4-7.2 in 2012, 7.0% CI95%:6.4-7.2 in 2017) [14, 27]. Other UCHP remained stable from 2007 to 2017.

Across Europe, about 40% of those questioned reported usage of UCHPs at least once in the preceding year over the same timeframe, but the various methods used make any comparisons across countries difficult [28, 29].

Concerning the pandemic’s impact, in 2021 [23], 6% of respondents started using UCHPs the year prior (i.e., since the pandemic’s onset), and 7% reported their UCHP usage had increased due to pandemic-related stresses. Conversely, 72% claimed their UCHP use had not changed, and 22% said their usage had “probably” altered due to the pandemic. In 2023, a relatively low percentage of respondents (between 2.2% for prayer, and 14.1% for essential oils) reported recent use of UCHPs within the last three years (i.e., since the pandemic, Table 1).

**Table 1.** Recalculation of data from the Harris (2019) [22] and the Odoxa (2023) [24] surveys on the reported use of UCHP.

|  | Harris survey (2019) |  |  | Odoxa survey (2023) |  |  |  |
| --- | --- | --- | --- | --- | --- | --- | --- |
|  | Frequency of UCHP use at least once in the past year among users<sup>a</sup> | Frequency of UCHP use at least once (lifetime)<sup>b</sup> | Frequency of UCHP use at least once in the past year (whole sample)<sup>c</sup> | Frequency of UCHP use at least once in the past year among users<sup>a</sup> | Frequency of UCHP use at least once (lifetime)<sup>b</sup> | Frequency of UCHP use at least once in the past year (whole sample)<sup>c</sup> | Frequency of UCHP use since < 3 years (pandemic)<sup>d</sup> |
| Acupuncture | 40.0% | 28.0% | 11.2% | 52.0% | 21.0% | 10.9% | 12.0% |
| Bach flowers |  |  |  | 90.0% | 13.0% | 11.7% | 44.0% |
| Chiropractic | 52.0% | 12.0% | 6.2% |  |  |  |  |
| Essential oils |  |  |  | 89.0% | 37.0% | 32.9% | 38.0% |
| Dietetician/fasting | 53.0% | 21.0% | 11.1% | 87.0% | 9.0% | 7.8% | 46.0% |
| Homeopathy | 54.0% | 33.0% | 17.8% | 73.0% | 42.0% | 30.7% | 19.0% |
| Hypnosis | 50.0% | 9.0% | 4.5% | 68.0% | 15.0% | 10.2% | 37.0% |
| Kinesiology |  |  |  | 75.0% | 10.0% | 7.5% | 31.0% |
| Magnetism |  |  |  | 70.0% | 16.0% | 11.2% | 31.0% |
| Meditation |  |  |  | 92.0% | 14.0% | 12.9% | 44.0% |
| Naturopathy | 71.0% | 10.0% | 7.1% |  |  |  |  |
| Osteopathy | 65.0% | 49.0% | 31.9% | 78.0% | 46.0% | 35.9% | 25.0% |
| Phytotherapy |  |  |  | 85.0% | 11.0% | 9.4% | 26.0% |
| Prayer |  |  |  | 94.0% | 8.0% | 7.5% | 27.0% |
| Psychology | 44.0% | 26.0% | 11.4% |  |  |  |  |
| Sophropogy | 55.0% | 14.0% | 7.7% | 64.0% | 15.0% | 9.6% | 40.0% |
| Yoga |  |  |  | 85.0% | 13.0% | 11.1% | 45.0% |

Note: UCHP: Unconventional Healthcare Practices; a = data from slide 23 of the Harris survey report [22], from slide 32 of the Odoxa survey report [24]; b = data from slide 22 (Harris), from slide 17 (Odoxa); c = a x b; d = data from slide 33 (Odoxa), data not available for Harris.
The frequency of UCHP use at least once in the past year is the data usually presented in the previous French IFOP survey [21]. This frequency was not directly available in the Harris and Odoxa surveys and was calculated (see column c) by multiplying the lifetime use of single UCHP (a) by the use of this single UCHP in the past year (b).

Overall, while survey comparisons should be approached with caution, these data do not support claims of an “explosion” or “exponential increase” in UCHP usage. Only small increases were observed in Switzerland, where a regulatory policy was implemented in 2010.

It is worth mentioning that several surveys mentioned an increase in the use of UCHP [23] although their results were in contradiction with that claim [22–24]. For instance in France, isolated percentages were overinterpreted as reflecting a diachronic trend [24] or previous data collected by the same survey company were simply ignored [22, 23].

#### 3.2. A surge of referrals in UCHPs-related misconducts?

In France, Miviludes’ data indicates an increase of 115.1%, in the number of health-related referrals between 2010 and 2017, followed by relative stability up to 2021 (see Figure 1A). Over the same period, the total number of referrals (any cause) escalated by 92.9%, with a particular increase in the proportion of security-related referrals on the total number of referrals between 2017 and 2021 (see Figure 1A and Table S2: 34.0% (621/1825) in 2010, 20.8% (537/2581) in 2017, and 37.7% (1515/4020) in 2021).

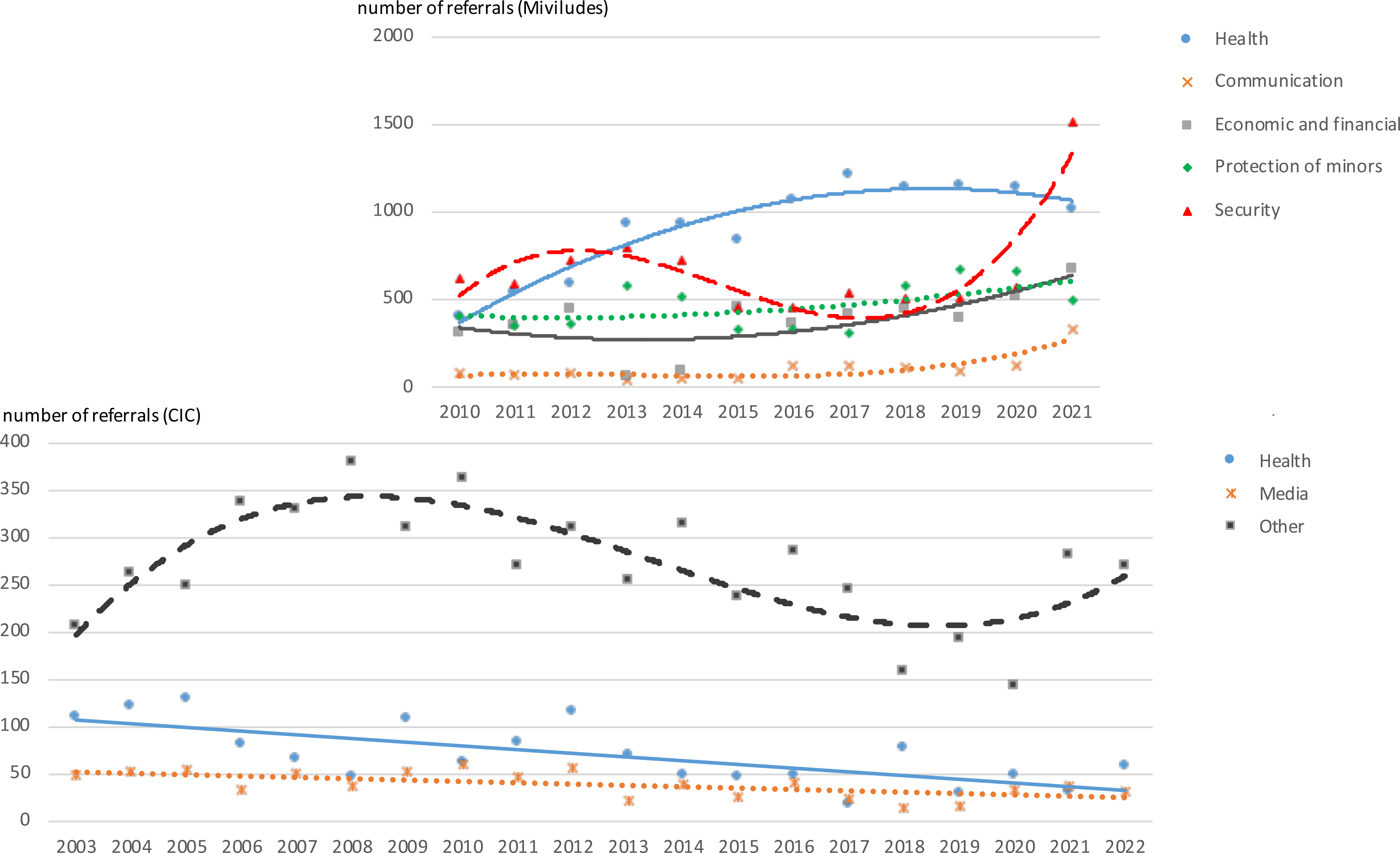

In contrast, the proportion of health-related referrals on the total number of referrals, increased from 22.0% (402/1825) in 2010 to 46.8% (1208/2581) in 2017, and then declined to 25.1% (1011/4020) in 2021 (refer to Table S2 in SM). Notably, the proportion of referrals relating to UCHP remained relatively stable (between 12 and 14%; see Table S2).

In terms of relative increases, Table 2 shows that from the five years preceding the pandemic to 2021, the number of health-related referrals remained fairly steady (+5.8%) compared to a 79.3% rise in overall referrals. The evolution from the pre-pandemic period to 2021 (post-pandemic), shows a slight decrease in the number of health-related referrals (− 11.7%), despite a 44.0% rise in total referrals.

**Table 2.** Relative increase (or decrease) of referrals and articles about UCHPs over time.

|  | Evolution from |  |  | Evolution from |  |  |
| --- | --- | --- | --- | --- | --- | --- |
|  | 2011 | 2016 | 2019 | 2011 | 2016 | 2019 |
|  | to 2021 |  |  | to last available data |  |  |
| Miviludes (referrals) |  |  |  | *last available data 2021* |  |  |
| Total | 92.9% | 79.3% | 44.0% |  |  |  |
| Health | 115.1% | 5.8% | -11.7% |  |  |  |
| Communication | 350.6% | 315.4% | 235.3% |  |  |  |
| Security | 151.2% | 234.2% | 198.5% |  |  |  |
| Economic/financial | 102.6% | 64.7% | 60.6% |  |  |  |
| Protection of minors | 31.0% | 51.9% | -20.9% |  |  |  |
| CIC (referrals) |  |  |  | *last available data 2022* |  |  |
| Total | -20.5% | 2.9% | 44.0% | -18.7% | 5.3% | 47.2% |
| Health | -56.6% | -33.4% | -40.3% | -21.1% | 21.1% | 8.5% |
| Communication | -29.0% | 15.2% | 145.2% | -40.2% | -3.0% | 106.5% |
| Other | -10.8% | 8.0% | 60.5% | -14.6% | 3.4% | 53.7% |
| Media (articles) |  |  |  | *last available data 2023* |  |  |
| Acupuncture | -32.6% | 1.7% | -27.7% | 66.3% | 150.8% | 78.3% |
| Homeopathy | 100.0% | 71.8% | -61.3% | 9.6% | -5.9% | -78.8% |
| Naturopathy | 516.0% | 396.8% | 81.2% | 1004.0% | 790.3% | 224.7% |
| Osteopathy | 45.7% | 6.3% | -3.6% | 100.0% | 46.0% | 32.4% |
| Miviludes | 136.0% | 223.1% | 141.4% | 187.6% | 293.8% | 194.3% |
| All excepted Miviludes | 77.1% | 64.1% | -27.8% | 146.6% | 128.6% | 0.6% |
| Pubmed (articles) |  |  |  | *last available data 2023* |  |  |
| Overall | 60.2% | 33.4% | 26.2% | 18.4% | -1.4% | -6.7% |
| France | 450.0% | 77.4% | 4.8% | 460.0% | 80.6% | 6.7% |
| Switzerland | 217.6% | 17.4% | -12.9% | 347.1% | 65.2% | 22.6% |

Note: UCHP: Unconventional Healthcare Practices; "Communication" refers to call for communications by the media or institutions; "Security" includes referrals related to the security of the nation (e.g., religious extremism, ecological terrorism, etc...); "Economic and financial" includes referrals of financial misconducts linked to sectarian deviances; "Protection of minors" includes referrals related to sectarian deviances representing a threat for minors

Comparing the last available data to that of five years before the pandemic (2016) or just before the pandemic (2019), the most significant increases were observed in two domains: security (+234.2% and +198.5%, respectively) and communication (+315.4% and +235.3%, respectively). The former includes referrals related to the security of the nation (e.g., religious extremism, ecological terrorism, etc…), the latter refers to call for communications by the media or institutions.

In Switzerland, the number of referrals received by the CIC reveals a gradual increase between 2003 and 2008-2009, followed by a decline until 2020, and subsequently an increase post-pandemic (see Figure 1B). Referrals related to health show a linear decrease from 2010 to 2022 (−21.1%). However, comparing the last available data to that of five years before the pandemic (2016) or just before the pandemic (2019), a slight uptick was observed (+21.1% and +8.5%, respectively; see Figure 1B and Table 2), probably due to the small number of those kind of referrals. In contrast, the overall number of referrals increased more clearly (+5.3% and +47.2%, respectively; see Figure 1B and Table 2). Communication-related referrals showed a linear decrease from 2010 to 2022 (−40.2%), but a relative increase post-pandemic (+106.5%). This was also probably due to the small number of those referrals and the rise observed in Switzerland was much smaller than that in France.

#### 3.3. An “explosion” of UCHP mentions in Media and a steady increase in Scientific Literature?

Analysis 1: From 2017 to 2023, the number of articles significantly increased for all topics, except for homeopathy, which saw a peak of occurrence only in 2018-2019, corresponding to the delisting of homeopathic medicines in France (see Figure 2A). Comparing the last available data to that of five years before the pandemic (2016) or just before the pandemic (2019), the most pronounced increases were observed for “Miviludes” (+293.8% and +194.3%, respectively) and “naturopathy” (+790.3% and +224.7%, respectively) (see Table 2).
Analysis 2: In contrast to media coverage, the number of scientific articles on UCHP showed a steady increase in international publications throughout the period studied (see Figure 2B and Table 2). However, comparing the last available data to that of five years before the pandemic (2016) or just before the pandemic (2019), a consistent increase of publications on this topic was found from teams located in both France (+80.6% and +6.7%, respectively) and Switzerland (+65.2% and +22.6%, respectively).

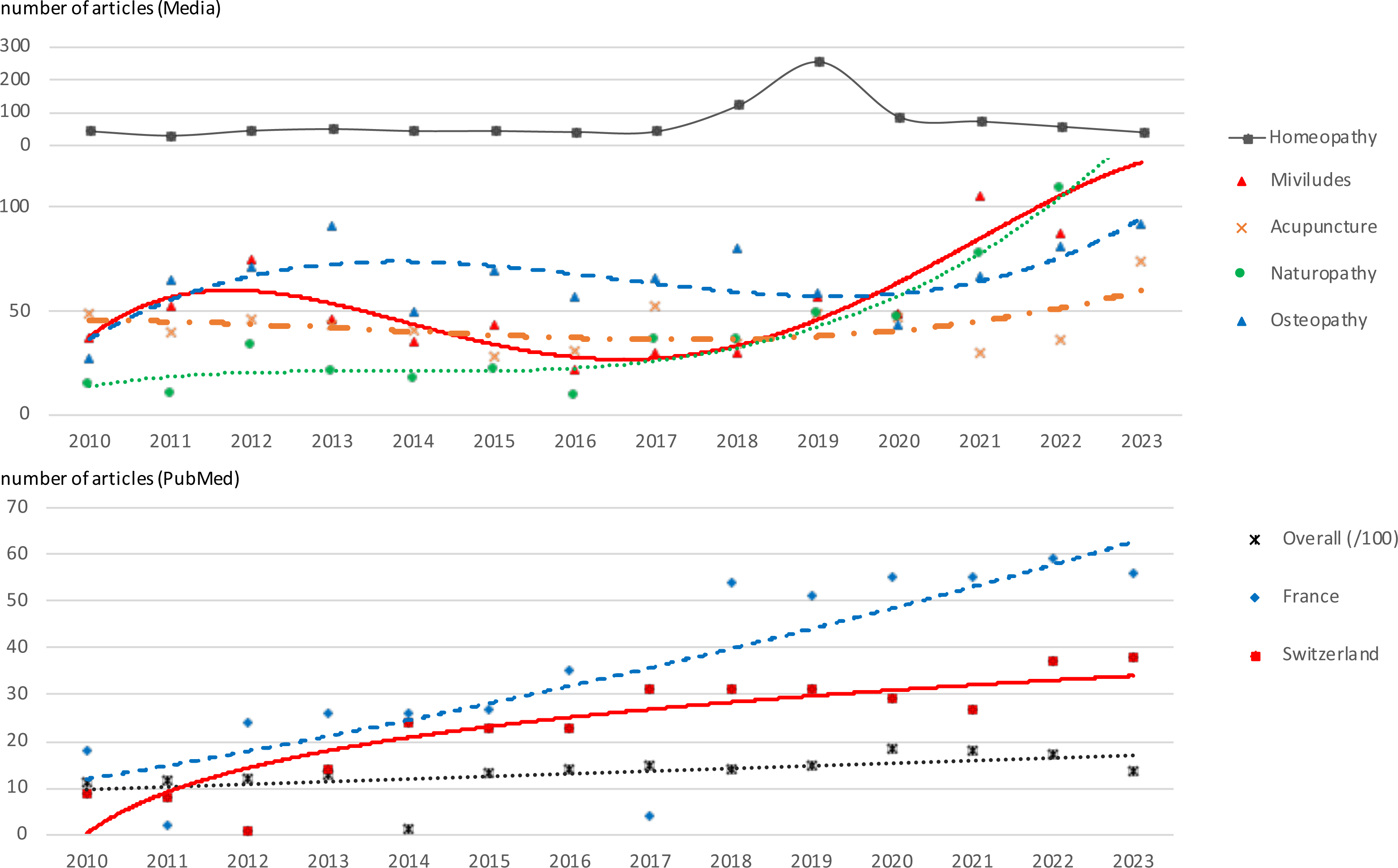

Data presented in Table 2 thus reveals a discrepancy between the moderate and consistent increase in scientific literature and the more rapid rise in the number of media articles (in particular related to Miviludes, Naturopathy and Acupuncture).

### 4. Discussion

The primary objective of this article was to rigorously assess any evidence of the purported “exponential increase” in the use of UCHP in France, which is frequently asserted by media outlets and certain official organizations, without proper references. Our comprehensive analysis of available information sources does not suggest an “explosion” in UCHP use or related sectarian misconduct, yet does show a steady increase in the number of mentions in media of UCHP.

In fact, surveys conducted in France since 2007 demonstrate a fairly stable reported use of UCHP by about 40%. It’s important to note that the most recent survey conducted in France (April 2023, [24]) focused specifically on “alternative medicine”, making it impossible to estimate which proportion of UCHP were utilized instead of conventional treatments or sometimes in conjunction with them [24]. This distinction is vital in accurately assessing UCHP-related risk, which is particularly significant when UCHP substitutes more effective conventional treatments [8]. Moreover, when asked about combining complementary and conventional treatments with an integrative perspective, 54 to 55% of those questioned in 2007 and 2019 reported using UCHP [21, 22]; and 88% and 76% considered UCHP as complementary to conventional treatments in 2019 and 2023 [22, 24]. In a different survey conducted in Switzerland in 2022, 79% of respondents expressed a preference for an “integrative approach” to healthcare, emphasizing the combination of UCHP with conventional medicine [30]. Interestingly, UCHP usage in Switzerland, where UCHP regulation was enacted in 2010, has only slightly increased (from 24.0 to 28.9% between 2007 and 2017). Hence, there seems to be little evolution of the population’s self-reported use. One needs to also consider that some evolutions in self-reports could be due to “changing willingness to disclose consumption” of UCHP rather than “actual change in use”.

Indeed, health-related referrals registered by Miviludes showed a steady increase (+115.1%) from 2010 to 2021, nearly parallel to the overall rise in referrals (+92.9%). However, during the five years preceding the pandemic, both the number and proportion of health-related referrals either remained stable or saw consistent decline. Obviously, reports to the instances require a set of facts and circumstances, and might only represent the tip of the iceberg of problematic behaviors. Nevertheless, the presented numbers call into question the concept of “constant increase” and “threat” presented by Miviludes and the media in the French context. Furthermore, in Switzerland, the CIC has noted a consistent linear decline in health-related referrals since 2010, unaffected by the pandemic.

Our analyses highlight that the terms “explosion” or “exponential increase” are more applicable to media representation of UCHP, particularly in relation to Miviludes’ media presence in France. This is evident from the sharp increase in media communications, press inquiries, and newspaper articles (see Figure 2A and Table S1). The above-mentioned Miviludes’ activity surge in the media sector illustrates this too (Figure 2A and Table S1). A similar trend, albeit to a much lesser extent, is observed for the CIC in Switzerland.

Our review of the different data available in the public sector underscore that the media-biased “polyphonic” epidemiology, of multiple sources perpetuating reports are not substantiated by facts. A critical analysis of the data is therefore crucial before publicizing them in media outlets or, more importantly, official reports [5, 11, 31]. Vigilance is required especially as some media appear increasingly influenced by UCHP skeptics [32, 33], some of whom represent vocal groups actively opposing UCHP use and spread [34].

#### 4.1. Limitations

We must acknowledge limitations of our work.

Firstly, as reported above, comparisons between French surveys with various methods, definitions and samples, require caution. By questioning evidence on a surge of UCHP in the French population, we should rather conclude that we have neither evidence to show an increase, nor evidence to strongly discard this hypothesis. In contrast, conclusions from the comparison of 3 sequential Swiss surveys are more reliable as they are based on a very large representative sample and use the same method [14].

Secondly, our media analysis included all articles featuring the aforementioned keywords, which doesn’t necessarily imply that every article provided an in-depth discussion of the UCHP in question. In fact, the delisting of homeopathy in 2018, the Covid-19 pandemic, and the increased focus on naturopathy due to the listing of naturopaths on the Doctolib platform (Doctolib is the main web platform dedicated to licensed health professionals in France) in 2022 have all contributed to an increase in critical UCHP analyses within newspaper articles over the past five years.

Thirdly, our research lacks a more qualitative assessment of how UCHPs were discussed in these media articles. For example, the tone could be factual, positive or critical. We acknowledge this aspect could be further explored.

Fourthly, we recognize that some organizations and media outlets, including Miviludes, claim an “explosion” in UCHP training offerings. This could suggest that a booming supply indicates a similarly booming demand. However, our study has not investigated the number of providers of UCHP, of training schools or of open practices. This could be important and helpful work to present in the future.

#### 4.2. Implications

The media’s heightened focus on UCHP and unsubstantiated claims in the past requires us to carefully scrutinize any future reported increases in UCHP usage or related misconducts. It is essential to differentiate between an actual rise in UCHP utilization or associated issues, and the impacts of increased media awareness. The latter might influence individuals to more readily report their use of UCHP or any potential abuses, for instance.

Interestingly, the most recent survey included in this review [24] reveals a perception gap: respondents believe that “the general French public” would be more likely to turn to UCHP compared to five years ago than they themselves would (70% vs. 54%). This difference between personal choices and the perceived behavior of the general population might suggest that those surveyed do not see themselves as part of the much-publicized “explosion” in UCHP usage.

When it comes to the risk of sectarian misconduct, the same survey [24] indicates that approximately 70% of respondents recognize that UCHP usage can potentially be associated with aberrant practices. Although nearly 30% fail to identify this risk, it appears that public awareness initiatives are having an impact. It is also important to note that the risk of misconduct is not unique to UCHP. The COVID-19 pandemic has shown publicly that representatives of conventional medicine can also stray from their Hippocratic oath in [35].

### 5. Conclusion

Facing this media-skewed, polyphonic epidemiology, Official organizations such as the Ministry of Health or the French National Medical Council should cite existing objective data, and perhaps support research. The technical support committee for UCHP regulation, recently established in France under the guidance of the Minister Delegate for Territorial Organization and Health Professionals [3], will find it advantageous to ground its analyses on factual evidence rather than opinions or speculation. Health care specialists, journalists and politicians should become aware of biases concerning UCHPs and foster a more informed and balanced discourse regarding these practices.

### Declarations

#### Ethics approval and consent to participate

not applicable

#### Consent for publication

not applicable

#### Availability of data and materials

all material is provided in the manuscript

#### Authors’ contributions

F.B., L.B., F.P., J.N. designed the study; F.B. collected the data and provided the first analyses; F.B., C.B. and N.F. provided critical evaluation of the data; F.B. wrote the main manuscript text and prepared all figures and tables. All authors critically reviewed and altered the manuscript.

#### Acknowledgements

## Data Availability

All data produced in the present work are contained in the manuscript

## Supplementary Material

**Table S1:**
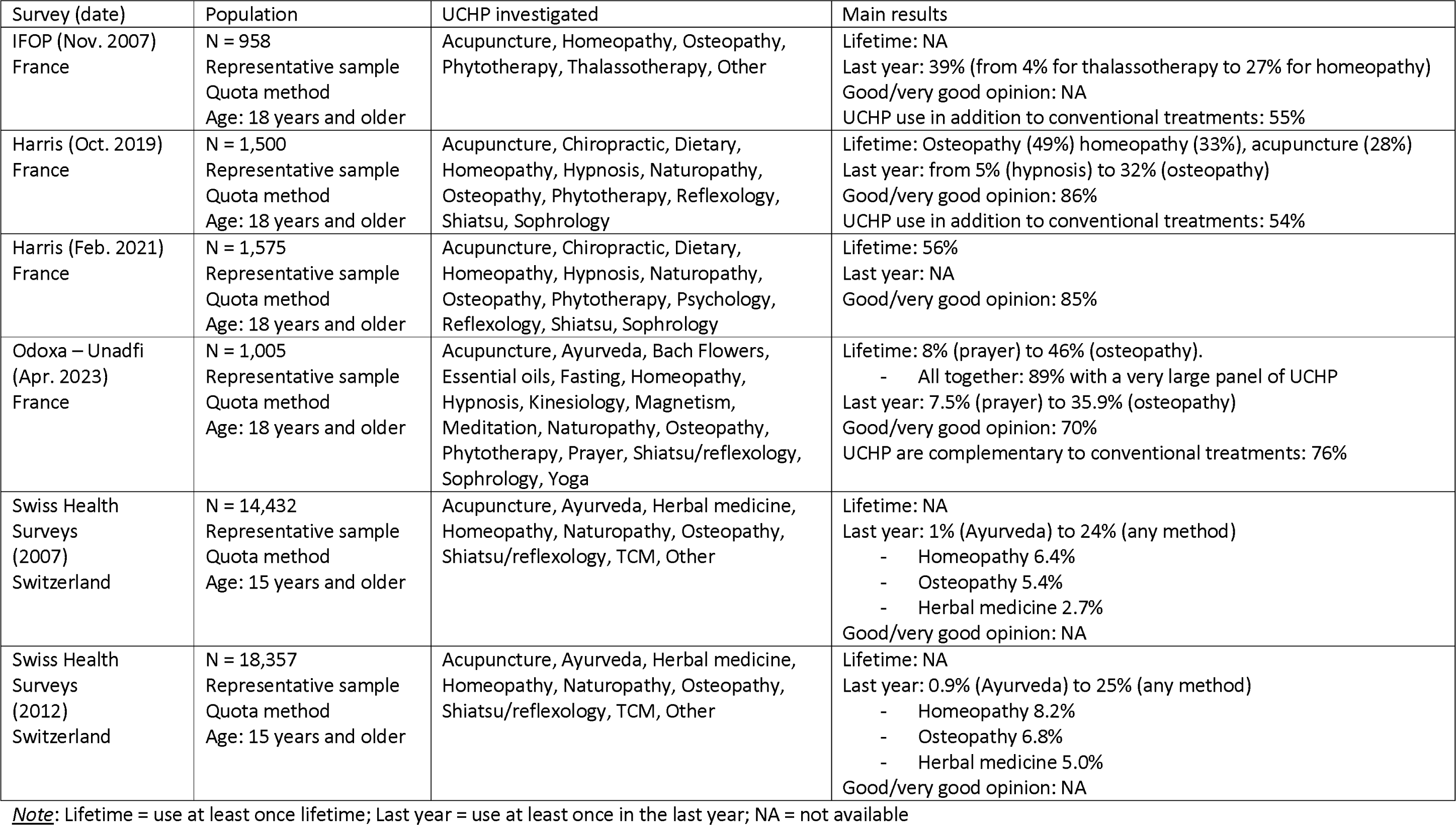
Summary results of the surveys conducted in France and Switzerland.

| Survey (date) | Population | UCHP investigated | Main results |
| --- | --- | --- | --- |
| IFOP (Nov. 2007)<br>France | N = 958<br>Representative sample<br>Quota method<br>Age: 18 years and older | Acupuncture, Homeopathy, Osteopathy,<br>Phytotherapy, Thalassotherapy, Other | Lifetime: NA<br>Last year: 39% (from 4% for thalassotherapy to 27% for homeopathy)<br>Good/very good opinion: NA<br>UCHP use in addition to conventional treatments: 55% |
| Harris (Oct. 2019)<br>France | N = 1,500<br>Representative sample<br>Quota method<br>Age: 18 years and older | Acupuncture, Chiropractic, Dietary,<br>Homeopathy, Hypnosis, Naturopathy,<br>Osteopathy, Phytotherapy, Reflexology,<br>Shiatsu, Sophrology | Lifetime: Osteopathy (49%) homeopathy (33%), acupuncture (28%)<br>Last year: from 5% (hypnosis) to 32% (osteopathy)<br>Good/very good opinion: 86%<br>UCHP use in addition to conventional treatments: 54% |
| Harris (Feb. 2021)<br>France | N = 1,575<br>Representative sample<br>Quota method<br>Age: 18 years and older | Acupuncture, Chiropractic, Dietary,<br>Homeopathy, Hypnosis, Naturopathy,<br>Osteopathy, Phytotherapy, Psychology,<br>Reflexology, Shiatsu, Sophrology | Lifetime: 56%<br>Last year: NA<br>Good/very good opinion: 85% |
| Odoxa – Unadfi<br>(Apr. 2023)<br>France | N = 1,005<br>Representative sample<br>Quota method<br>Age: 18 years and older | Acupuncture, Ayurveda, Bach Flowers,<br>Essential oils, Fasting, Homeopathy,<br>Hypnosis, Kinesiology, Magnetism,<br>Meditation, Naturopathy, Osteopathy,<br>Phytotherapy, Prayer, Shiatsu/reflexology,<br>Sophrology, Yoga | Lifetime: 8% (prayer) to 46% (osteopathy).<br>- All together: 89% with a very large panel of UCHP<br>Last year: 7.5% (prayer) to 35.9% (osteopathy)<br>Good/very good opinion: 70%<br>UCHP are complementary to conventional treatments: 76% |
| Swiss Health<br>Surveys<br>(2007)<br>Switzerland | N = 14,432<br>Representative sample<br>Quota method<br>Age: 15 years and older | Acupuncture, Ayurveda, Herbal medicine,<br>Homeopathy, Naturopathy, Osteopathy,<br>Shiatsu/reflexology, TCM, Other | Lifetime: NA<br>Last year: 1% (Ayurveda) to 24% (any method)<br>- Homeopathy 6.4%<br>- Osteopathy 5.4%<br>- Herbal medicine 2.7%<br>Good/very good opinion: NA |
| Swiss Health<br>Surveys<br>(2012)<br>Switzerland | N = 18,357<br>Representative sample<br>Quota method<br>Age: 15 years and older | Acupuncture, Ayurveda, Herbal medicine,<br>Homeopathy, Naturopathy, Osteopathy,<br>Shiatsu/reflexology, TCM, Other | Lifetime: NA<br>Last year: 0.9% (Ayurveda) to 25% (any method)<br>- Homeopathy 8.2%<br>- Osteopathy 6.8%<br>- Herbal medicine 5.0%<br>Good/very good opinion: NA |

Note: Lifetime = use at least once lifetime; Last year = use at least once in the last year; NA = not available

**Table S2:**
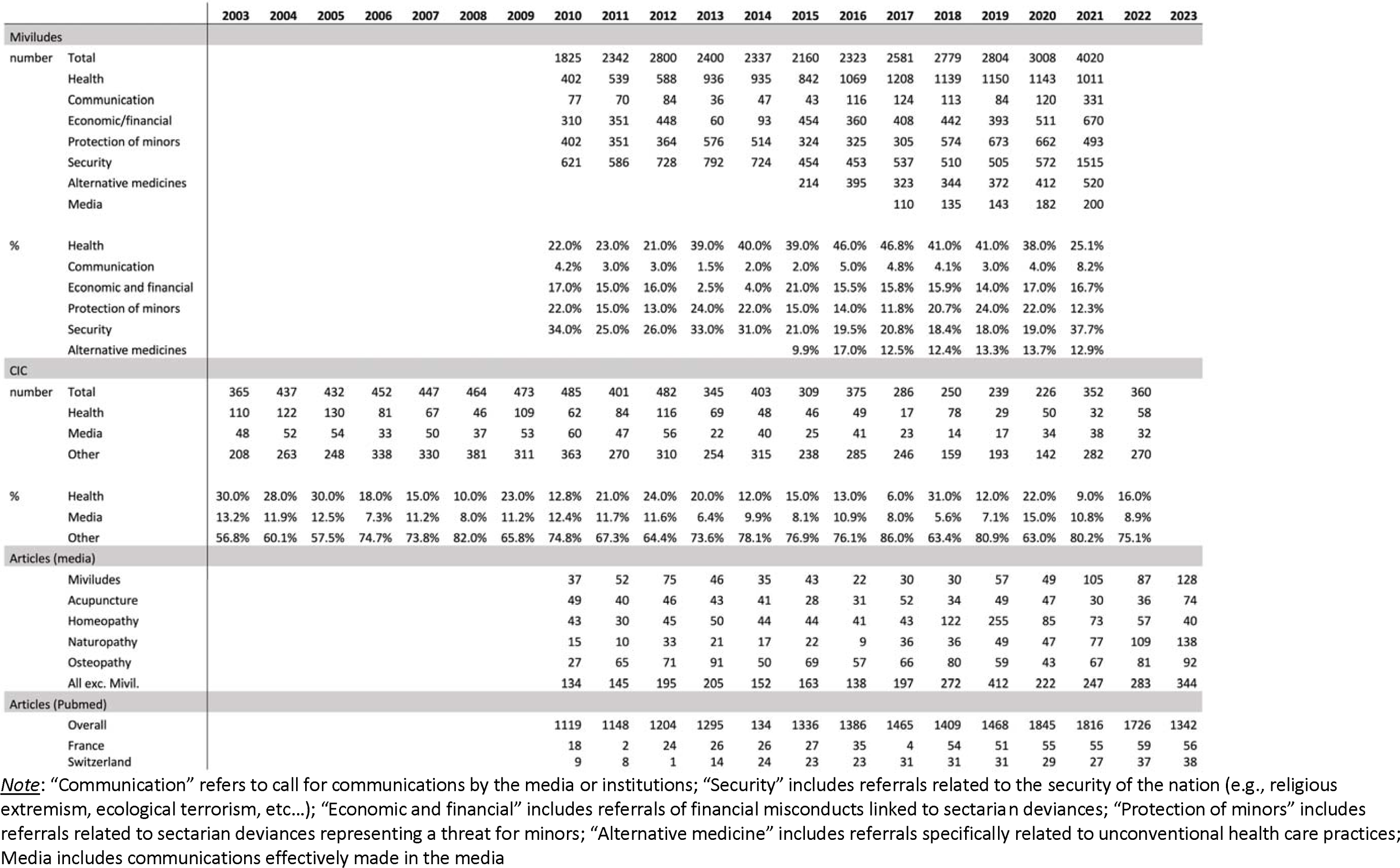
Number and percentage of referrals or articles from 2003 to 2023.

